# Estimation of the onset ratio and the number of asymptomatic patients of COVID-19 from the proportion of untraceable patients

**DOI:** 10.1101/2021.07.28.21261241

**Authors:** Takashi Odagaki

## Abstract

A simple method is devised to estimate the onset ratio of COVID-19 patients from the proportion of untraceable patients tested positive, which allows us to obtain the number of asymptomatic patients, the number of infectious patients and the effective reproduction number. The recent data in Tokyo indicate that there are about six to ten times as many infectious patients in the city as the daily confirmed new cases. It is shown that, besides social distancing and use of effective masks, a quarantine measure on non-symptomatic patients is critically important in controlling the pandemic.

## Introduction

The pandemic COVID-19 is still prevalent in most parts of the world despite of continuous efforts by governments to control it. Difficulty of the control lies in the fact that patients of COVID-19 take a route different from common epidemics. In common epidemics, infected individuals show symptoms and become infectious after an incubation period. Then they are treated and recover from the disease. In COVID-19, infected individuals become infectious before symptom-onset and asymptomatic patients who do not show any symptoms before their recovery are infectious. In order to formulate proper strategies in controlling COVID-19, it is important to know the proportion of asymptomatic and pre-symptomatic patients [1-3], who will be called non-symptomatic patients collectively, in addition to the number of symptomatic patients. However, it is unpractical to identify all non-symptomatic patients in the entire population by PCR tests. Therefore, it is an important problem to devise a method for estimating the number of non-symptomatic infectious patients from data reported daily such as the confirmed new cases and the proportion of untraceable patients tested positive. Here, symptomatic patients are divided into two groups; one is traceable patients who know from whom they have gotten infected and the other is untraceable patients who have had no contact with pre-symptomatic and symptomatic patients.

In this paper, I propose a simple method by which the onset ratio *x* of COVID-19 patients can be estimated from the proportion *f* of untraceable patients tested positive and show that the proportion of the infectious patients can be obtained from the proportion of untraceable patients. I first analyze the infection process on the basis of the SIQR (Susceptible-Infectious-Quarantined-Removed) model [4] and find a relation between *x* and *f*. Then, I argue that the number of infectious patients and the number of new patients on a given day can be related to the proportion *f*. I also discuss the effective reproduction number which depends on a quarantine rate of non-symptomatic patients and show that the quarantine of non-symptomatic patients is critically important in controlling COVID-19. I analyze the situation in Tokyo and discuss why COVID-19 does not converge in Tokyo.

### Classification of infectious patients

I classify infected individuals on the basis of the SIQR model [4] and the mean field approach as follows. I first assume that patients of COVID-19 follow the same disease progression day by day at the same pace on average since their infection. Namely, I assume that the timeline of infection follows infected, infectious, symptom/quarantined or asymptomatic and that the time between these events is deterministic. On a particular day which I call day zero, there are many infected individuals who can be classified by the number of days since their infection. The number of patients, some of whom are identified as patients by PCR test and quarantined on the day zero, is denoted by *n*_0_. I denote the average onset ratio of COVID-19 by *x*, and then the number of infected symptomatic individuals in the group of patients is given by *xn*_0_ and the number of asymptomatic individuals in the group of patients is given by (1 − *x*)*n*_0_. I denote by *n*_−1_, *n*_−2_, …, *n*_−*k*_ the number of infected individuals who got infected one, two,…, *k* days later than the day when the *n*_0_ individuals got infected. Similarly, I denote by *n*_1_, *n*_2_, …, *n*_*ℓ*−1_the number of individuals who got infected one, two, …, *ℓ* − 1 days earlier than the day when the *n*_0_individuals got infected [5]. Here, *k* is the sum of the infectious period before symptom-onset and the period between the onset date and the quarantined date, and *ℓ* denotes the infectious period of asymptomatic patients after symptomatic patients in the same group are quarantined. I assume that the proportion 1 − *x* of asymptomatic patients is common in every group of patients. Figure 1 shows schematically the breakdown of patients into these three groups, where patients in the shaded area are infectious. Note that in the present analysis, the latent and incubation periods do not play any roles.

**Fig. 1.**
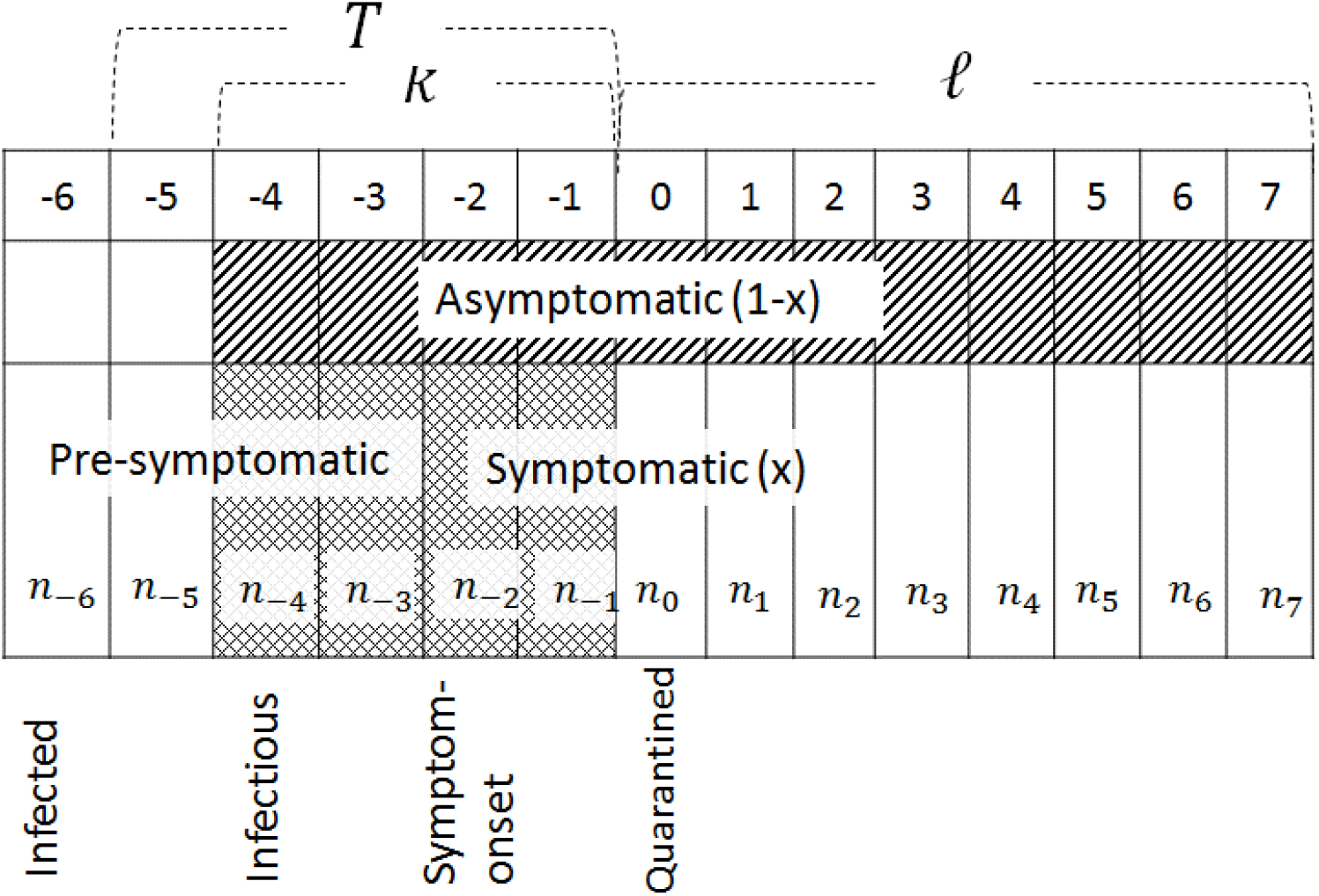
Break down of COVID-19 patients into asymptomatic, pre-symptomatic and symptomatic patients. Patients in the shaded area are infectious. The hatched region represents infectious asymptomatic patients who produce untraceable patients and the cross-hatched region corresponds to infectious pre-symptomatic and symptomatic patients who produce traceable patients. *T* denotes the effective period of PCR test before quarantine.

I assume that all symptomatic patients will be tested intentionally some days after the symptom-onset and quarantined. I also assume that PCR tests, which is effective since *T* days before day zero, are conducted on the general population and denote the quarantine rate of patients by *q*. If the quarantine measure on infected individuals is taken effectively, these infected patients decrease by a factor (1 − *q*) every day. Therefore, the number of infectious patients ℑ on day zero is given by

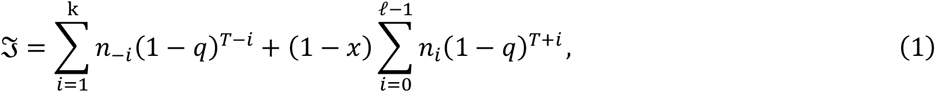

and the new patients Δ*I* infected on day zero who will be identified some days later are given by

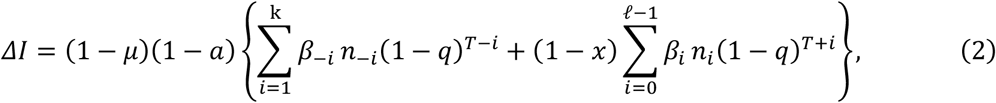

where *β*_*j*_ is the infection coefficient of patient group *n*_*j*_, *μ* is the fraction of immunized individuals who are no longer susceptible, and *a* represents the reduction ratio of social contacts among people due to lockdown measures. Here, I assume *T* ≥ *k* for simplicity.

In order to make the following description transparent, I define an average of *n*_*j*_ and a weighted average of *β*_*j*_ as follows: average of *n*_−1_, *n*_−2_, …, *n*_−*k*_

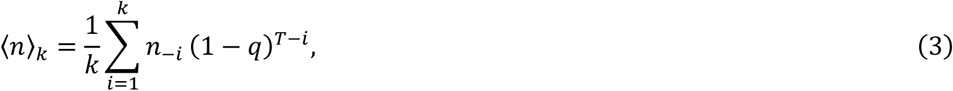

average of *n*_0_, *n*_1_, *n*_2_, …, *n*_*ℓ*−1_

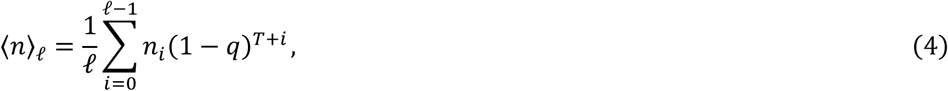

weighted average of *β*_−1_, *β*_−2_, …, *β*_−*k*_

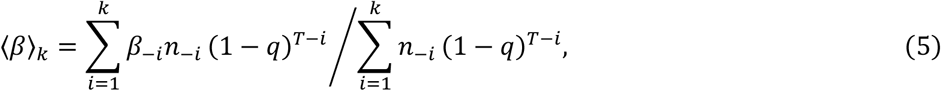

weighted average of *β*_0_, *β*_1_, *β*_2_, …, *β*_*ℓ*−1_

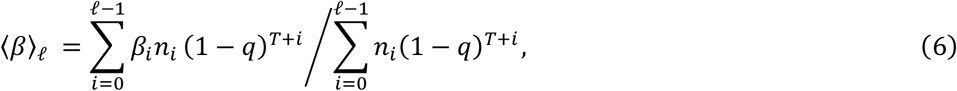

and write Eqs. (1) and (2) as

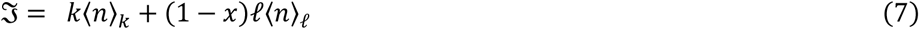

and

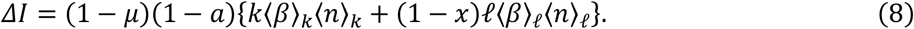

### Relation between the proportion of untraceable patients and the onset ratio

In this section, I argue that the traceability of patients can be related to infection from asymptomatic patients and derive a relation between the onset ratio and the proportion of untraceable patients. Out of the newly infected individuals Δ*I*, Δ*Q* ≡ *x*Δ*I* will show symptoms some days later and (1 − *x*)Δ*I* will not show any symptoms. Patients Δ*Q* showing symptoms will be identified by PCR tests and be listed as daily confirmed new cases. I assume that they are classified into two groups Δ*Q*_*t*_ and Δ*Q*_*unt*_, where Δ*Q*_*t*_are traceable patients who are infected from symptomatic and pre-symptomatic patients and Δ*Q*_*unt*_ are untraceable patients who are infected from asymptomatic patients. Namely, Δ*Q*_*t*_ and Δ*Q*_*unt*_ are given by

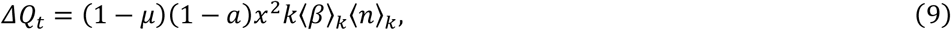

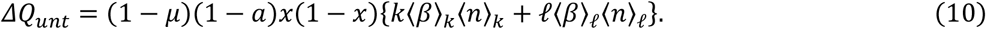

Therefore, the proportion of untraceable cases in the daily confirmed new cases is given by

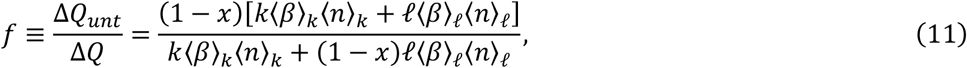

which does not depend on *μ* and *a* explicitly. This relation can be inverted to get

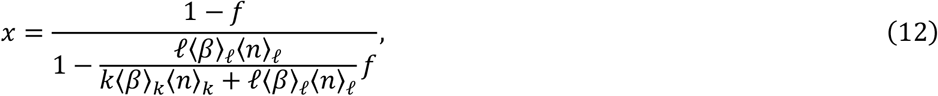

which indicates that the onset ratio *x* can be obtained from the proportion of untraceable patients *f* once other parameters are known.

In order to incorporate the trend of infection status [6] into the present analysis, I first define trend parameters *τ*_*k*_ and *τ*_*ℓ*_by *τ*_*k*_ = ⟨*n*⟩_*k*_/(1 − *q*)^*T*^*n*_0_ and *τ*_*ℓ*_ = ⟨*n*⟩_*ℓ*_/(1 − *q*)^*T*^*n*_0_. It is apparent that the infection status after day zero will be increasing, stationary and decreasing when *τ*_*k*_ *>* 1, *τ*_*k*_*=* 1 and *τ*_*k*_ *<* 1, respectively. Similarly, *τ*_*ℓ*_ > 1, *τ*_*ℓ*_ = 1 and *τ*_*ℓ*_ *<* 1 indicate that the infection status before day zero has been decreasing, stationary and increasing, respectively. It is straight forward to express *x* in terms of *τ*_0_ and *τ*_1_ as

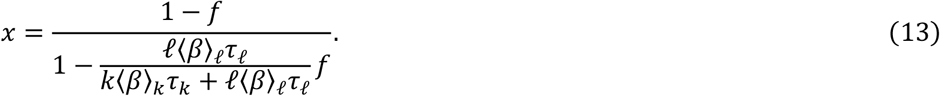

Therefore, Δ*I* is written as

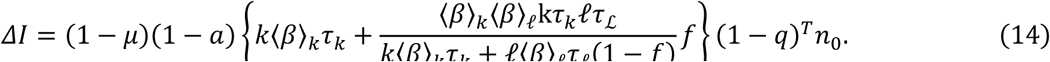

Since the confirmed new cases on day zero (Δ*Q*)_0_ is given by (Δ*Q*)_0_ = *x*(1 − *q*)^*T*^*n*_0_,ℑ can be expressed in terms of *τ*_*k*_, *τ*_*ℓ*_ and (Δ*Q*)_0_ as

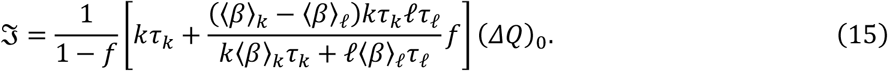

### Number of infectious patients and asymptomatic patients in Tokyo

As an example of analysis, I apply the present analysis to the infection status in Tokyo in the middle of June, 2021. In Tokyo, PCR tests have not conducted on non-symptomatic patients and I can set *q* = 0. I assume that the infection status was stationary in the middle of June, 2021, and set *τ*_*k*_ = *τ*_*ℓ*_ = 1 and ⟨*β*⟩_*k*_ = ⟨*β*⟩_*ℓ*_. Then, Eq. (13) reduces to

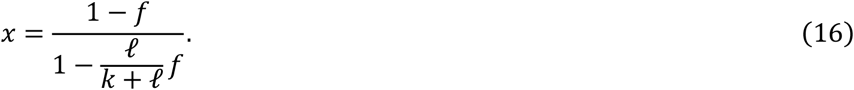

According to Ministry of Health, Labour and Welfare of Japan (MHLW), transmission occurs in the 2 days before symptom-onset and in the 7∼10 days post symptom-onset [7]. These estimations are consistent with those reported in Ontario, Canada: Ontario Agency for Health Protection and Promotion reported that the transmission occurs in the 3∼5 days before onset and in the 8∼10 days post onset [5]. Assuming on average a patient will be tested and quarantined next day of symptom-onset, I set as a model case *k* = *3days* and *ℓ*= *7 days*. Then, Eq. (16) is reducible to

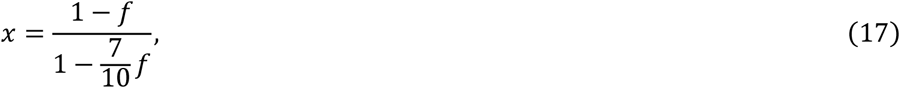

which is shown in Fig.2. In Tokyo, the proportion of untraceable patients is *f* = 50∼60% [8]. Using the lower value *f* = 50 %, I find *x* = 77%. This value becomes *x* = 69% if *f* = 60% is used. These values are consistent with observations of the onset ratio *x* = 76% [9] or 75% [10].

**Fig. 2.**
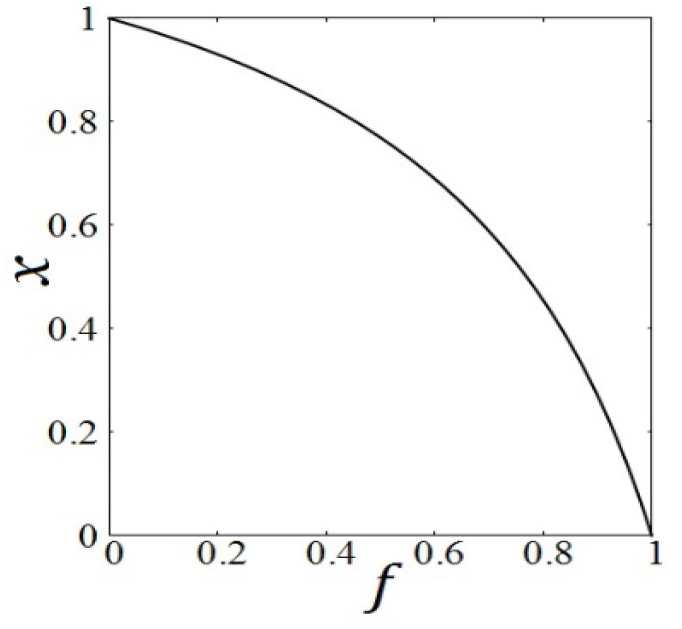
The onset ratio *x* is plotted as a function of the proportion of untraceable patients *f* when *k* = 3 days and *ℓ = 7 days*.

On the same conditions, the number of infectious patients Eq. (15) can be written as

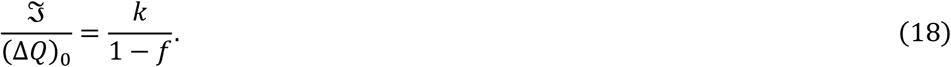

If there were no untraceable patients, all infected individuals would be symptomatic and, therefore, the number of infectious patients will be given by the number of new cases times days during which they are infectious, i.e. ℑ/(Δ*Q*)_0_ *= k* when *f =* 0 as Eq. (18) indicates. Equation (18) shows ℑ/(Δ*Q*)_0_ = 2*k* and 2.5*k* when *f* = 50 ∼60 % respectively. Figure 3 shows *f* dependence of ℑ/(Δ*Q*)_0_ when *k* = *3days* and *ℓ* = *7days*. It is important to note that in Tokyo *f* = 50 ∼60% and *k* = *3days*, and thus there are 6∼7.5 times more infectious patients than the daily confirmed new cases. It should also be mentioned that the number of asymptomatic infectious patients ℑ_*as*_ excluding pre-symptomatic patients is given by

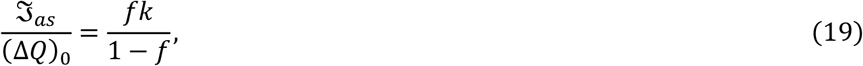

and thus ℑ_*as*_/(Δ*Q*)_0_ = 3 when *k* = *3days, ℓ* = *7days* and *f* = 50%.

**Fig. 3.**
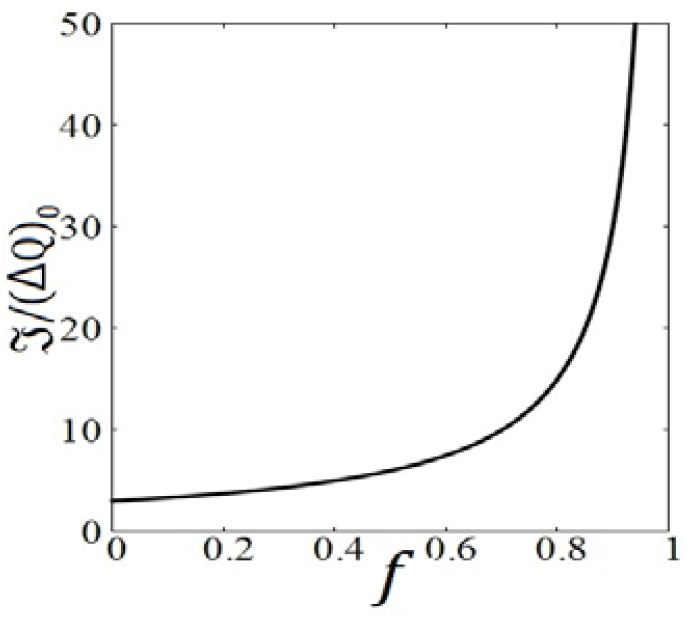
The ratio of the number of infectious patients in the city ℑ and the daily confirmed new cases (Δ*Q*)_0_ is plotted as a function of the proportion of untraceable patients *f* when *k* = *3days*.

In January 2022, the number of patients in Tokyo was increasing due to the omicron variant. I can assume that the difference between ⟨*β*⟩_*k*_and ⟨*β*⟩_*ℓ*_ is small and *q* is negligible in Eq. (15). From the infection curve increasing exponentially with rate 0.2, *τ*_*k*_is estimated to be *τ*_*k*_ *=* 1.2*4*. In this period, the proportion of untraceable patients was 65.5% [8], and thus, setting *k* = *3days*, I get ℑ/(Δ*Q*)_0_ ≈ 11.

Around the end of February 2022, the sixth wave in Tokyo [8] seems to be in the stationary state with *f* = 61%, and thus I can set *τ*_*k*_ = 1. Therefore, ℑ/(Δ*Q*)_0_ is estimated to be ℑ/(Δ*Q*)_0_ ≈ 7.7.

### Effective reproduction number and assessment of policies

I first define an effective infectious period *d*_*eff*_ and an effective infection coefficient *β*_*eff*_ as follows:

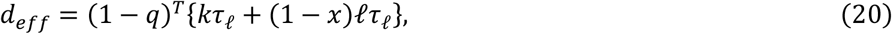

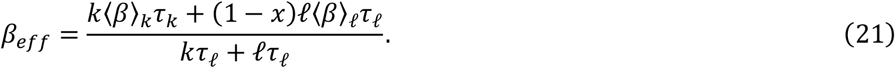

Equation (8) can now be expressed as

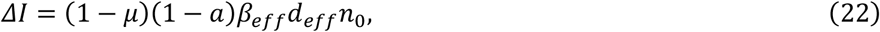

and the effective reproduction number *R*_*eff*_ on day zero is given by

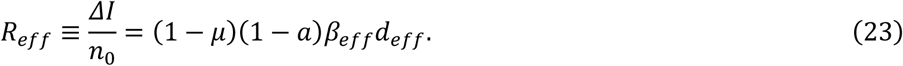

It should be remarked that when *d*_*eff*_ is equal to the recovery time or the inverse of the recovery rate *γ* and *β*_*eff*_ is a constant, the expression for *R*_*eff*_ is identical to that defined in the standard SIR model. It is important to note that, in contrast to the standard SIR model, the effective reproduction number *R*_*eff*_ defined by Eq. (23) depends on the quarantine rate of non-symptomatic patients through *d*_*eff*_, which has a significant effect in reducing the effective reproduction number.

The basic strategy against COVID-19 is to bring the effective reproduction number smaller than unity so that the number of patients decreases. Equation (23) indicates that there are three tactics: (1) increase immunized people by vaccination, if it is effective in preventing transmission of SARS-CoV-2, or by natural immunity to make (1 − *μ*) smaller, (2) enforce social distancing by various lockdown measures to make (1 − *a*) smaller, and (3) quarantine asymptomatic patients by PCR tests to make *d*_*eff*_ smaller. In countries like Taiwan, Australia and New Zeeland who have succeeded in controlling COVID-19 before the vaccination was started, PCR tests have been conducted more than 100 times per positive patient in a well-designed manner.

In Tokyo, the PCR test has been used only to confirm the infection of SARS-CoV-2 for people who show some symptoms, which means *q* ≈ 0, and thus it has not been contributing to the battle against COVID-19. Furthermore, lockdown measures have been very sloppy. Instead of enforcing social distancing among the entire population it has been applied only specific targets, like the night life district on 2020 and restaurants serving alcoholic beverages in 2021 and 2022. The policy targeting on certain shops and opening hours has only limited effects on social distancing since people gather together in parks or on streets. Furthermore, the policy has been enforced and lifted every one or two months, which has caused the wavy infection curve [11,12]. It could be possible to increase *a* by, for example, promoting Telework, limiting working days, reducing crowd in commuter trains and banning gatherings.

## Discussion

In the present analysis, traceable and untraceable patients tested positive are related to infection from pre-symptomatic and symptomatic patients and from asymptomatic patients, respectively. Although this assumption may not be rigorous, it is a good assumption if people cooperate in the investigation of infection route by the health department of local government.

When the infection status is increasing or decreasing [6], the relation Eq. (15) between the number of infectious patients and the number of the newly confirmed new cases reduces to

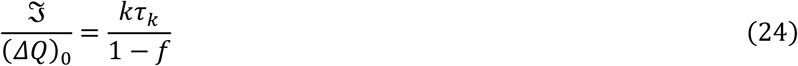

if the condition ⟨*β*⟩_*k*_ *=* ⟨*β*⟩_*ℓ*_ is satisfied. This ratio becomes large in the increasing status and small in the decreasing status compared to Eq. (18). In this case, the effective reproduction number depends on the infection status through *d*_*eff*_. In general, *d*_*eff*_ is an increasing function of *τ*_*k*_ and *τ*_*ℓ*_. Since the increasing state corresponds to *τ*_*k*_ *>* 1 *> τ*_*ℓ*_ and the decreasing state to *τ*_*k*_ *<* 1 *< τ*_*ℓ*1_, *d*_*eff*_ can increase or decrease depending on the relation between *τ*_*k*_ and *τ*_*ℓ*_.

The infection coefficient of SARS-CoV-2 depends on variants. When a new variant with stronger infection coefficient emerges, ⟨*β*⟩_*k*_becomes larger than ⟨*β*⟩_*ℓ*_. The number of infectious patients will increases for a stronger variant when other parameters are the same.

In 2021, vaccination has been progressed in many countries and the infection status seems to be improving at least in the reduction of serious cases. The effect of vaccination appears through *μ* in Eq. (2) which depends on variants of the virus. Therefore policies for controlling COVID-19 should not rely only on the vaccination, and a proper combination of policies on vaccination, social distancing and quarantine of non-symptomatic patients must be designed in each country around the world [13].

## Conclusion

It has been shown that the onset ratio and the number of infectious patients can be estimated from the proportion of untraceable patients tested positive. The effective reproduction number depends not only on the vaccination rate and effects of social distancing but also on *d*_*eff*_ which is controlled by quarantine measures on pre-symptomatic and asymptomatic patients. The effect of the quarantine measure appears as reduction of the effective reproduction number by (1 − *q*)^*T*^ which could be significantly large compared to the effects of the lockdown measures and the vaccination.

## Data Availability

N/A

## Acknowledgments

This work was supported in part by JSPS KAKENHI Grant Number 18K03573.

## References

1. D. P. Oran and E.J, Topol, Prevalence of Asymptomatic SARS-CoV-2 Infection. Ann. Intern. Med. 174, 286–287 (2021). https://doi.org/10.7326/L20-1285.

2. R. Subramanian, Q. He, and M. Pascual, Quantifying asymptomatic infection and transmission of COVID-19 in New York City using observed cases, serology, and testing capacity, Proc. Nat. Acad. Sci., 118, e2019716118 (2021). https://doi.org/10.1073/pnas.2019716118

3. B. T. Montague, M. F. Wipperman, A. T. Hooper, S. C. Hamon, R. Crow, F. Elemo, L. Hersh, S. Langdon, J. D. Hamilton, M. P. O’Brien, E.A.F. Simões. Anti-SARS-CoV-2 IgA Identifies Asymptomatic Infection in First Responders. J. Infect. Dis. 225, 578–586 (2022). https://doi.org/10.1093/infdis/jiab524

4. T. Odagaki, Analysis of the outbreak of COVID-19 in Japan by SIQR model, Infectious Disease Modelling, 5, 691–698 (2020). https://doi.org/10.1016/j.idm.2020.08.013

5. Ontario Agency for Health Protection and Promotion (Public Health Ontario). COVID-19 overview of the period of communicability – what we know so far. Toronto, ON: Queen’s Printer for Ontario; 2021. https://www.publichealthontario.ca/-/media/documents/ncov/covid-wwksf/2021/03/wwksf-period-of-communicability-overview.pdf?la=en

6. T. Odagaki and R. Suda, Classification of the infection status of COVID-19 in 190 countries, J. Clinical Trials 11, 472 (2021). https://doi.org/10.1101/2020.12.17.20248445

7. Ministry of Labour, Health and Welfare, Japan, “Guidance for treatment of COVID-19” ver5.1, July, 5, (2021). https://www.mhlw.go.jp/content/000801626.pdf

8. Tokyo Metropolitan Government, “Updates on COVID-19 in Tokyo”, July 20 (2021) https://www.fukushihoken.metro.tokyo.lg.jp/iryo/kansen/corona_portal/info/monitoring.images/0715graph3-1.JPG

9. M. A. Johansson, T. M. Quandelacy, S. Kada, P. V. Prasad, M. Steele et al, SARS-CoV-2 Transmission From People Without COVID-19 Symptoms, JAMA Network Open. 4(1) e2035057 (2021). http://doi:10.1001/jamanetworkopen.2020.35057

10. M. Alene, L. Yismaw, M. A. Assemie. D. B. Ketema, B. Mengist et al, Magnitude of asymptomatic COVID-19 cases throughout the course of infection: A systematic review and meta-analysis, PLoS ONE 16(3), e0249090 (2021). https://doi.org/10.1371/journal.pone.0249090

11. T. Odagaki, Self-organized wavy infection curve of COVID-19, Scientific Reports, 11, 1936 (2021). https://doi.org/10.1038/s41598-021-81521-z

12. T. Odagaki, Self-organization of oscillation in an epidemic model for COVID-19, Physica A 573, 125925 (2021). https://doi.org/10.1016/j.physa.2021.125925

13. A. Singh, M. Arquam, Epidemiological modeling for COVID-19 spread in India with the effect of testing, Physica A: Statistical Mechanics and its Applications 592, 126774 (2022). https://doi.org/10.1016/j.physa.2021.126774

